# Estimating the spreading and dominance of SARS-CoV-2 VOC 202012/01 (lineage B.1.1.7) across Europe

**DOI:** 10.1101/2021.02.22.21252235

**Authors:** Nicolò Gozzi, Matteo Chinazzi, Jessica T. Davis, Kunpeng Mu, Ana Pastore y Piontti, Marco Ajelli, Nicola Perra, Alessandro Vespignani

## Abstract

We develop a two strain, age-structured, compartmental model to assess the spreading potential of the B.1.1.7 variant across several European metropolitan areas and countries. The model accounts for B.1.1.7 introductions from the UK and different locations, as well as local mitigation policies in the time period 2020/09 − 2021/02. In the case of an increase of transmissibility of 50%, the B.1.1.7 variant has the potential to become dominant in all investigated areas by the end of March 2021.

---

A new variant of SARS-CoV-2, called Variant Of Concern (VOC) – 202012/01 and often referred to as lineage B.1.1.7 [1], was discovered in the United Kingdom in mid September [2, 3]. Mutations have been reported and tracked since the beginning of the outbreak [4]. However, the emergence of the B.1.1.7 lineage has been related to a steep increase in the number of cases in the UK [1, 5], triggering strict non-pharmaceutical interventions such as lockdowns (see England and Scotland), quarantines, and travel bans from passengers traveling from the area in more than 40 countries.

Here, we estimate the spreading of the new variant in metropolitan areas across Denmark, Germany, Greece, Italy, Poland, Portugal, and Spain. We use a stochastic, age-structured, two strain epidemic model that considers the introduction of the B.1.1.7 variant from the UK and other locations as estimated from travel and mobility flows [6–8]. We model the dynamic of the B.1.1.7 variant and the wild type considering mitigation policies and find that, in the case of a 50% increase in transmissibility, the new variant has the potential to become the dominant strain in all European areas studied by the end of March 2021. We estimate that in all locations analyzed the probability that the B.1.1.7 variant is not able to establish local transmission is less than 1%. In the locations studied, vaccination campaigns are not projected to affect a large fraction of the population in the first quarter of 2021 and the model does not account for them. The results suggest great caution in planning the relaxation of non-pharmaceutical interventions as we approach spring and call for the necessity of strengthening genomic surveillance efforts.

## Results

We adopt an age-structured, two strain, compartmental model calibrated on the evolution of confirmed deaths, separately, for each geographical region studied. We initialize the model by using the prevalence and arrivals of the new variant estimated via a global stochastic, spatial, and age-structured metapopulation data-driven model [6–8]. Here, we report the results for major metropolitan areas in Germany (Berlin and Frankfurt) Italy (Milan and Rome), and Spain (Barcelona and Madrid). Due to lack of specific data about confirmed deaths for some of these municipalities, we extend our analysis to their administrative regions. Specifically, we consider the European NUTS2 (NUTS1 for Germany) territories. We also include in our analysis a set of specific countries, namely Denmark, Greece, Poland and Portugal. We initialize individuals into ten age groups [0−9, 10−19, 20−24, 25−29, 30−39, 40− 49, 50−59, 60−69, 70−79, 80+], considering official data of resident populations on January 1^*st*^, 2020, in the aforementioned areas.

The model assumes the emergence of the B.1.1.7 variant in the UK in the week 38 of 2020 (2020/09/13-2020/09/19), with an effective transmissibility that is 30% to 70% times that of the wild type. The model is calibrated on the weekly incident deaths recorded in each administrative region from 2020/09/01 to 2021/02/14 by using an Approximate Bayesian Computation (ABC) technique [9]. The model accounts for non-pharmaceutical interventions (NPIs) implemented by different governments and local authorities up to 2021/02/14 by incorporating data from Google’s COVID-19 Community Mobility Reports [10] and the Oxford COVID-19 Government Response Tracker [11] (see Supplementary Information).

We run the calibrated model between 2020/09/01 and 2021/03/31 including daily importations of the new variant in each region projected by the Global Epidemic and Mobility Model (GLEAM, see the Supplementary Material for more details). We then monitor the fraction of new daily cases attributable to the new variant in the investigated regions. As a baseline case, we assume that the B.1.1.7 variant has an increase in transmissibility of 50%. In order to account for NPIs after week 6, 2021 (2021/02/08- 2021/02/14), we have considered three scenarios:

- `Status quo`: NPIs and population behaviors remain the same of those observed in week 6, 2021.
- `Conservative relaxation:` contacts at work and in community setting are raised by 25% with respect to week 6, 2021. Furthermore, we also consider a conservative relaxation of measures targeting schools (equivalent to 1 step in the Oxford COVID-19 Government Response Tracker index on week 6, 2021 [11]).
- `Moderate relaxation:` this scenario assumes a 50% increase of contacts at work and in the community with respect to week 6, 2021. For schools, we consider a relaxation of two levels (i.e., subtracting 2) to the Oxford COVID-19 Government Response Tracker index on week 6, 2021 [11].

In Figure 1 we report the results concerning the estimated share of new infections caused by the B.1.1.7 variant over time, from 2021/01/01 through 2021/03/31. The projections (median, 90% CI) of the percentage of cases caused by the B.1.1.7 variant on 2021/03/31 are summarized in Table 1. Values vary from a median of 74% for Frankfurt, in the `Status Quo` scenario, to over 90% in Barcelona, Madrid, Portugal, and Denmark. Figure 1 shows that in our projections the new variant becomes dominant by the end of March 2021. In fact, in all cases, its share crosses the dashed horizontal line representing the 50% threshold. Barcelona, Denmark, Berlin, Madrid, Greece, and Portugal intersect the dashed line in February. In Milan, Frankfurt, Poland, and Rome, our projections indicate a share of the new variant above 50% in the first half of March. In Table 2 we report the median projected week (with 90% CI) in which the B.1.1.7 variant is expected to become dominant in different regions and restriction scenarios. The table confirms that, across all the cases considered in Europe, VOC 202012/01 is expected to become dominant before mid-March.

**Table 1:**
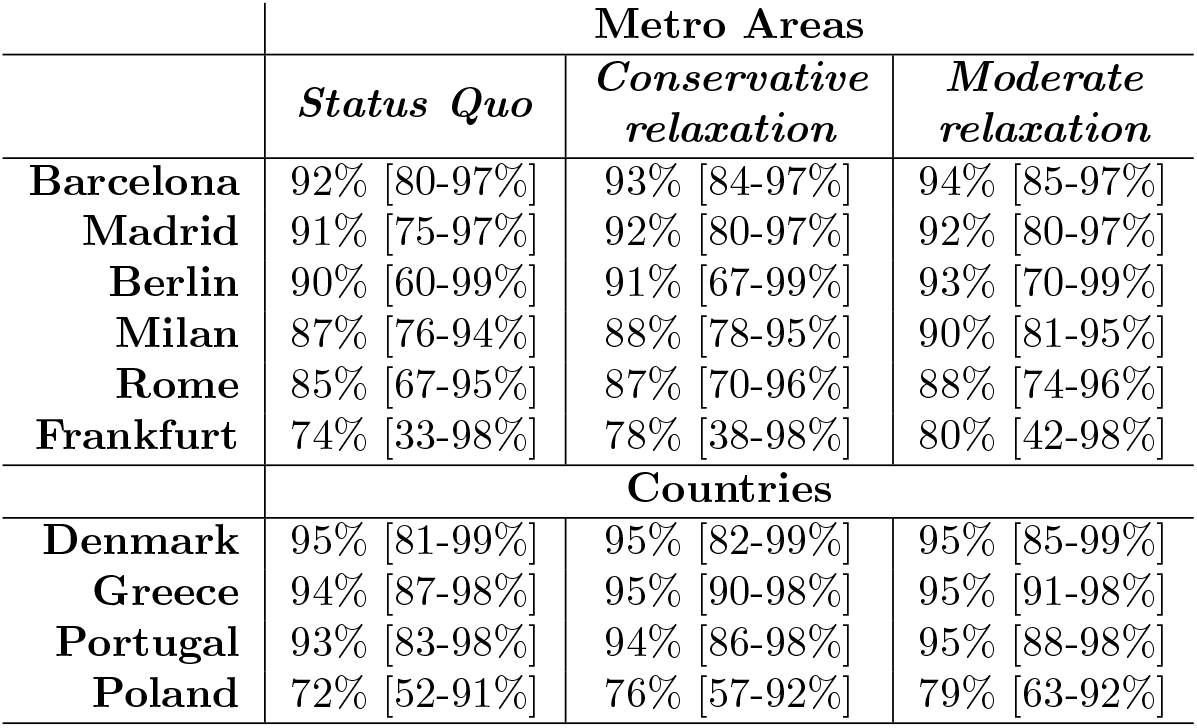
Percentage of new cases attributable to the B.1.1.7 variant on 2021/03/31. We summarize (median and 90% CI) the results for different regions and scenarios. We consider a variant transmissibility increase of 50%.

**Table 2:** Projected week of dominance of VOC 202012/01. We summarize (median and 90% CI) the projected week of dominance of the B.1.1.7 variant (i.e. 50% of new cases are attributable to the variant) for different regions and restrictions scenarios. We consider a variant transmissibility increase of 50%. “/” indicates that the projected week of dominance is beyond our simulation horizon (2021/03/31).

|  | Metro Areas |  |  |
| --- | --- | --- | --- |
|  | *Status Quo* | *Conservative relaxation* | *Moderate relaxation* |
| **Barcelona** | 8 [6-10] | 8 [5-10] | 8 [5-10] |
| **Madrid** | 8 [5-11] | 8 [5-10] | 8 [5-10] |
| **Berlin** | 8 [1-12] | 8 [2-12] | 8 [1-11] |
| **Milan** | 9 [7-11] | 9 [7-11] | 9 [7-10] |
| **Rome** | 9 [6-12] | 9 [6-11] | 9 [6-11] |
| **Frankfurt** | 11 [4-/] | 11 [4-/] | 10 [4-/] |
|  | Countries |  |  |
|  | *Status Quo* | *Conservative relaxation* | *Moderate relaxation* |
| **Denmark** | 7 [1-10] | 7 [1-10] | 7 [1-10] |
| **Greece** | 7 [4-9] | 7 [4-9] | 7 [3-8] |
| **Portugal** | 7 [3-9] | 7 [3-9] | 7 [3-9] |
| **Poland** | 11 [8-13] | 11 [8-13] | 11 [8-12] |

**Figure 1:**
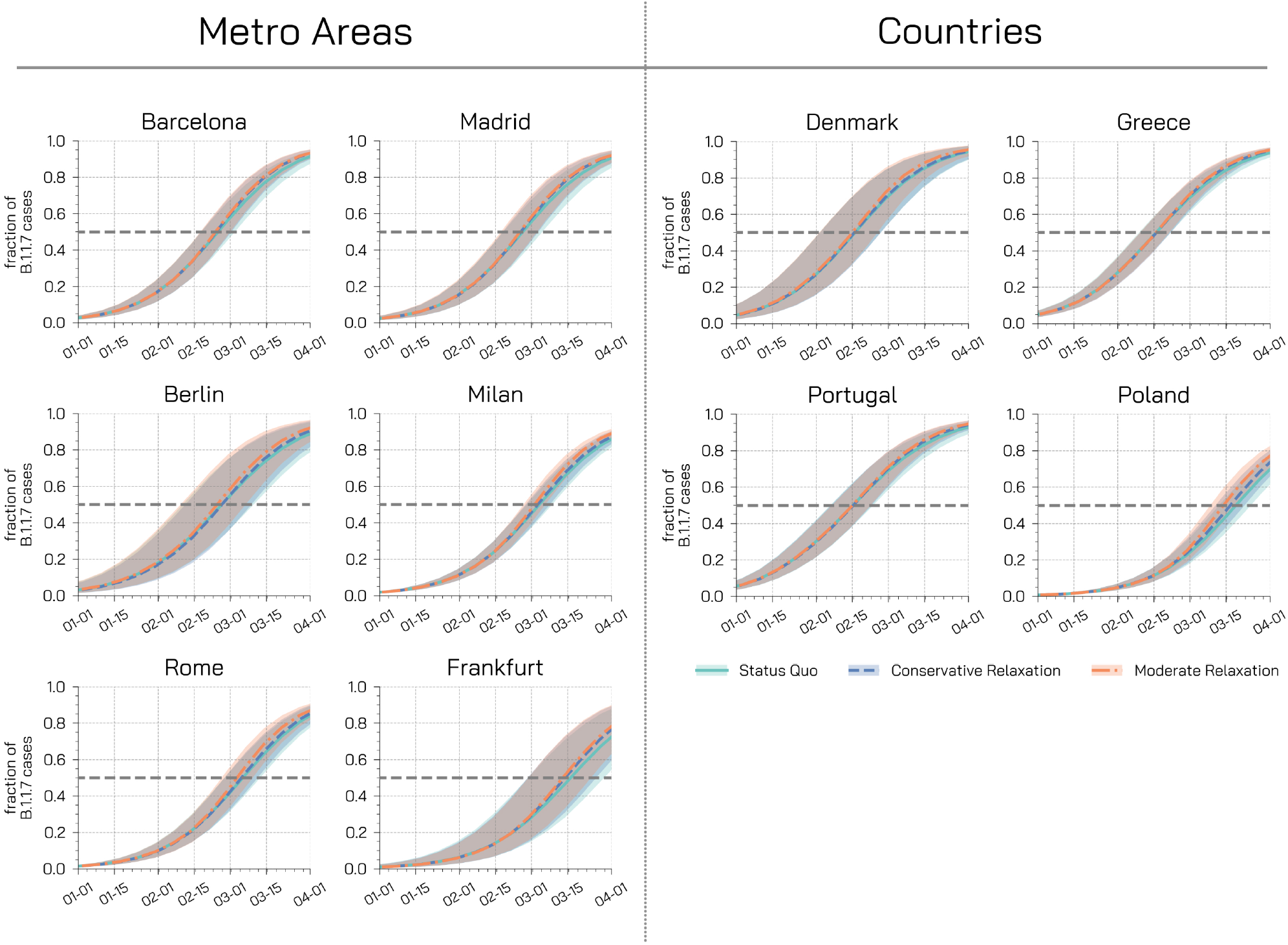
Fraction of new cases attributable to the B.1.1.7 variant. Each plot shows the fraction of new weekly cases attributable to the variant in different regions for different restrictions scenarios under the assumption of a 50% increase in transmissibility. Dashed horizontal lines represent the dominance threshold of 50% of new cases caused by the B.1.1.7 variant. The shaded areas represent the 50% CI.

We have also estimated the probability that B.1.1.7 is not able to establish local transmission in the different regions by measuring the fraction of runs that show a zero incidence of the variant in the last week of observation. These are the runs where a macroscopic outbreak of the new variant did not take off despite the multiple introduction events. In all cases considered, this fraction is smaller than 1%, hinting to a small chance that the variant is not already spreading locally in the considered administrative regions.

In most countries wide-scale genome sequencing capabilities are still lacking, making it difficult to obtain quantitative estimates of B.1.1.7 presence as a function of time. Nonetheless, recent estimates of B.1.1.7 dissemination in Denmark, Italy, and Portugal are available thanks to the analysis of nationwide RT-PCR Spike gene drop out data [12]. In Figure 2 we compare the model estimates (median, 50% CI, 95% CI) of the percentage of new cases attributable to the B.1.1.7 variant with the estimate from the Danish [13], Italian [14] and Portuguese [12] genomic data. It is worth stressing that the flash survey on B.1.1.7 prevalence in Italy was carried out at the country level, and disaggregated data are not available to date. In the Supplementary Material we also present sensitivity analyses considering an increase of 30% and 70% in the transmissibility of the new variant. The overall picture is confirmed, but of course the smaller/higher increase of transmissibility induces a slower/faster growth in the share of the new variant, respectively.

**Figure 2:**
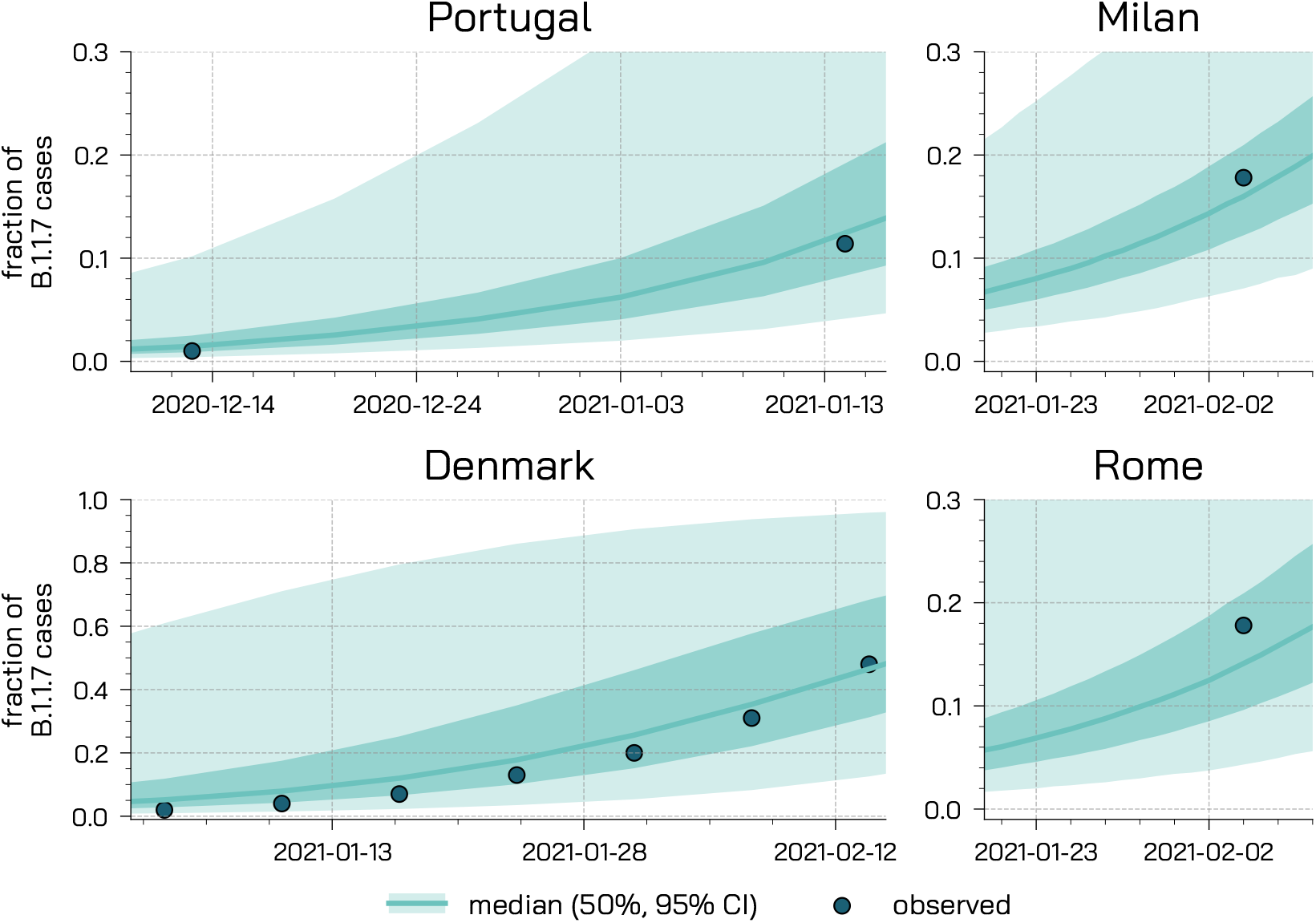
Model projections and real data. We compare the model projections (median, 50%, 95% CI) of the percentage of new cases attributable to VOC 202012/01 with observed data in Portugal, Denmark, and Italian regions (Milan and Rome).

## Limitations

We consider a simple transmission model that neglects the differentiation between symptomatic, pre- symtomatic, and asymptomatic transmission. The model does not account for spatial heterogeneity in adoption of NPIs, population density, infection rates, and mobility within each one of administrative regions considered. We have assumed that, except for the transmissibility, the new variant is characterized by the same key parameters as the wild type. However, it is important to note that some preliminary research studies point to the fact that the B.1.1.7 variant might lead to worst health outcomes and higher mortality [15]. Considering the short time horizon of our projections, we have neglected the potential impact of the vaccine rollout. In line with the current plans and response strategies, the scenarios analyzed assume either a status quo or a relaxation of NPIs and behaviors over the next three months. Hence, we neglect possible reversion in these policies in response to future spikes of new cases. The introductions of the B.1.1.7 variant in the various regions estimated by GLEAM does not consider the detection of asymptomatic/pre-symptomatic individuals due to testing requirements (i.e., COVID-free flights, international travel policies). Furthermore, GLEAM assumes independent case introductions neglecting the possibility of correlated arrivals due, for example, to family travel. Finally, while GLEAM is accounting for national and international mobility restrictions, the probability of traveling is considered to be independent of the risk of exposure of the individuals.

## Data Availability

See supplementary information for all details

## Discussion

The B.1.1.7 variant was able to become the dominant strain in several parts of England in about three months [16]. Initial analysis of genomic data suggested a growth rate 71% (95%*CI* : 67% − 75%) higher than other wild type SARS-CoV-2 lineages [5]. Public Health England estimated that an area with an effective reproductive number *R*_*t*_ = 0.8 (without the new variant) would have an *R*_*t*_ = 1.32 [95%CI 1.2 − 1.5] if only the new variant was spreading [16]. Detailed analysis of the secondary attack rates conducted using data from the National Health Service (NHS) in the UK Test and Trace program suggests an increase of 10% − 70% or 30% − 50% (depending on which type of data is used to select candidates of the new variant) with respect to the wild type across regions and age groups [16]. The results from a two strain epidemic model fit to the various NHS regions confirm this picture indicating the new variant as 56% more transmissible (95%*CI* : 50% − 74%) [17]. Preliminary analysis of the global spreading patterns of VOC 202012/01 suggest, as of December 20, 2020, multiple introductions in several countries, including France, Italy, Spain, and the USA among others [6, 18]. Many of such countries have since confirmed the detection of the B.1.1.7 variant in local cases with no travel history, despite travel bans and restrictions. Modeling results in the USA and France project that the new variant could become the dominant strain in those regions by March 2021 [19, 20]. Here, we contribute to this literature reporting on the spreading and dominance of the B.1.1.7 variant in several European regions using a two strain, age-structured, epidemic model. After calibrating the model during the period 2020/09/01 to 2021/02/14, we simulated the evolution of the variant and wild type SARS-CoV-2 through March 31, 2021. In order to account for mitigation policies after February 14, 2021 we considered a status quo scenario and two scenarios with different degrees of relaxation of the mitigation policies. Assuming the B.1.1.7 variant has an increased transmissibility of 50% with respect to the wild type, our simulations indicate probabilities of local extinctions, following importations of cases of the new variant, smaller than 1%. This suggests a likely onset of local transmission of the B.1.1.7 variant across all regions studied here. We estimate that by mid March the share of cases of the new variant, across all scenarios, will pass the 50% dominance threshold in the European regions and countries studied. While data concerning the prevalence of the new variant outside of the UK are scarce, our results appear in line with observations from Denmark [13], Italy [14], and Portugal [12].

In summary, our findings suggest that with high likelihood, sustained local transmission of the B.1.1.7 variant has started in all regions under investigation and highlight the importance of genomic surveillance to monitor the spread of this and other variants as well as the key role of NPIs in limiting the spread of this new variant as we move forward with the vaccine rollouts.

## Authors contributions

N.G., M.C., J.T.D., N.P. and A.V. designed the study. N.G. implemented the epidemic model, run and analyzed the simulations. M.C. implemented GLEAM model and run the simulations. M.C., J.T.D., K.M., and A.P.y.P. analyzed GLEAM simulations and provided the importation data from GLEAM. N.G., N.P., and A.V. wrote the first draft of the manuscript. All authors edited and approved the manuscript.

### Acknowledgements

MC, MA, and AV acknowledge support from COVID Supplement CDC-HHS-6U01IP001137-01. MC and AV. acknowledge support from Google Cloud and Google Cloud Research Credits program to fund this project. The findings and conclusions in this study are those of the authors and do not necessarily represent the official position of the funding agencies, the National Institutes of Health, or the U.S. Department of Health and Human Services.

## Supplementary Information

### Geographic regions studied

We consider the NUTS2 units (and NUTS1 for Berlin and Frankfurt) where the metropolitan areas are located: i) Berlin state (population 3.3 *M*) for Berlin, ii) Hessen (population 6*M*) Frankfurt, iii) Lombardy (population 10*M*) for Milan, iv) Latium (population 5.7*M*) for Rome, v) the Community of Madrid (population 6.7*M*) for Madrid, and vi) Catalonia for Barcelona (population 7.7*M*). Furthermore, considering the recent spike of cases and rapid increase of the B.1.1.7 variant’s share, we consider the country of Denmark (population 5.8*M*), Poland (population 37.8*M*), Portugal (population 10.2*M*), and Greece (population 10.4*M*)

### Demographic and epidemiological data

The census information is extracted from the `Italian National Institute of Statistics` (ISTAT) [21], the `Spanish Statistical Office` [22], *the* `Statistical Institute of Catalonia` [23], the German census [24], and the United Nations World Population Prospects [25]. For Latium, Lombardy, Hessen, Berlin, Denmark, Poland, Greece, and Portugal we use epidemiological data (daily COVID-19 deaths and cases) collected by the Johns Hopkins University Center for Systems Science and Engineering [26], while we use data from Ref. [27] for the Community of Madrid and Catalonia.

### Disease transmission model

The disease transmission is modeled with a SLIR compartmental scheme (see Figure 3). Susceptible and healthy individuals are placed in the *S* compartment. Interacting with infectious, *S* transit to the latent compartment (*L*). After the latent period *ϵ*^−1^, *L* individuals become infectious (*I*). Infectious eventually recover and transition to *R* after the infectious period *µ*^−1^. A fraction of these individuals unfortunately die. We consider the age-stratified Infection Fatality Rates (IFRs) from Ref. [28] and consider a delay of Δ days between the transition to the *R* compartment and actual death. This delay is in place to account for the time that elapses between the isolation of acute cases (i.e., hospitalization) and the official notification of death, which could span longer than two weeks [29]. We modify this general framework to include two virus strains. This is implemented considering two latent (*L*_1_, *L*_2_) and infectious (*I*_1_, *I*_2_) compartments. We assume that the two different strains have different transmission rates (*β*_1_ and *β*_2_) but the same latent and infectious period and IFRs. To account for the observations mentioned above, we assume that *β*_2_ = *β*_1_(1 + *ψ*) where *ψ* > 0 is the increase in transmissibility of the new variant. We consider individuals divided into 10 age groups and we define the number of contacts between age groups with a contacts matrix **C**. The element *C*_*ij*_ describes the average number of contacts that an individual in age group *i* has with individuals in *j* per day. It is important to note how the matrix **C** is split into four contributions that account for contacts at home, workplace, school, and other locations. We adopt country-specific contacts matrices developed in Ref. [30]. This compartmentalization setup has been previously used to study the interplay between virus strains in England [17].

**Figure 3:**
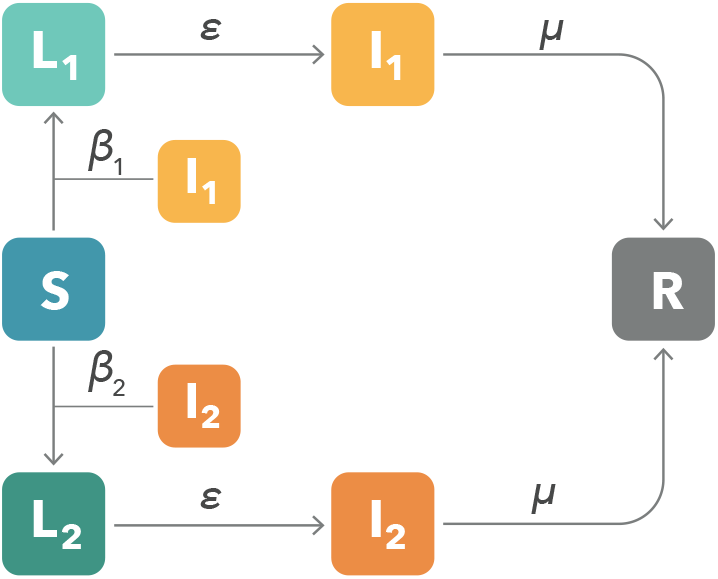
Compartmental structure. We adopt an extension of the classic SLIR model including a second strain. The transmission rate of the second strain is set as *β*_2_ = *β*_1_(1 + *ψ*). The structure is further extended to include 10 age brackets and an age-stratified contact matrix **C**. Finally, we account for age-stratified infection fatality rates (IFRs) to estimate the fraction of recovered that die after a delay of Δ days.

In addition, we consider a seasonal forcing as in Ref. [7] to account for differences in humidity, temperature, and other factors which might affect transmissibility, contact patterns, and ultimately *R*_*t*_ [31]. To this end, we rescale *R*_*t*_ → *s*_*i*_(*t*)*R*_*t*_ with the following function:

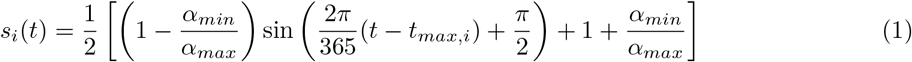

where *i* refers to the hemisphere considered. Note how all regions considered are located in the northern hemisphere. In the tropical region the scaling function is equal to 1. The value *t*_*max,i*_ is the time corresponding to the maximum of the sinusoidal function. It is fixed to January 15^*th*^ in the northern hemisphere and six months later in the southern one. We set *α*_*max*_ = 1 and consider *α*_*min*_ as free parameter (see more details below).

### Modeling of mitigation policies

We consider individuals divided into 10 age groups: [0 − 9, 10 − 19, 20 − 24, 25 − 29, 30 − 39, 40 − 49, 50 *−* 59, 60 − 69, 70 − 79, 80+]. As mentioned above, the contacts matrix **C** considers interactions in four specific social settings: contacts at school (**C**_*school*_), workplace (**C**_*work*_), home (**C**_*home*_), and in the general community (**C**_*community*_). Therefore, in general the contacts matrix is a linear combination of the four contributions according to the contacts reductions in different locations **C** =Σ^*s*^ *ω*_*s*_**C**_*s*_, where *ω*_*s*_ indicates the number of contacts per setting, and *s* indicates the different settings mentioned before. The baseline *ω*_*s*_ and **C**_*s*_ values for each specific country are from Ref. [30]. We quantify the time varying contacts reduction due to mitigation policies by using Google mobility reports [10] and at school using the Oxford Coronavirus Government Response Tracker [11]. We assume no changes to the number of contacts at home, though mitigation policies tend to increase their duration [32]. From the Google mobility reports we use the field workplaces percent change from baseline to infer contacts reduction in workplaces, the average of the fields retail and recreation percent change from baseline and transit stations percent change from baseline for the general community settings. The Google mobility report provides the percentage change *r*_*l*_(*t*) on day *t* of total visitors to specific locations *s* with respect to a pre-pandemic baseline. We turn this quantity into a rescaling factor for contacts such as *ω*_*s*_(*t*) = *ω*_*s*_(1+*r*_*l*_(*t*)/100)^2^, by considering that the number of potential contacts per location scales as the square of the the number of visitors. We also use the ordinal index C1 School closing from the Oxford Coronavirus Government Response Tracker to modulate contacts in schools and universities. The index ranges from a minimum of 0 (no measures) to a maximum of 3 (require closing all levels). Furthermore, all *ω* factors are multiplied (or set equal to in case of contacts at home) by setting-specific weights from Mistry et al. [30].

In order to simulate mitigation policies after the last collected data on week 6, 2021, we propose different scenarios regarding the contact reductions. The first scenario is based on a `status quo` situation in which we keep the contact matrices unchanged with respect to week 6 of 2021. The scenario simply assumes that NPIs and mobility behavior remain the same as those observed in week 6, 2021. We then propose two scenarios describing a `conservative` and a `moderate` relaxation of social distancing and other NPIs. In particular, in the conservative scenario contacts at work and in community setting are raised by 25% with respect to the observations in week 6, 2021. Furthermore, we also consider a conservative relaxation of measures in schools subtracting, if possible, 1 to the Oxford Coronavirus Government Response index on week 6, 2021 [11]. The third scenario describes a moderate relaxation of measures characterized by a 50% increase of contacts at work and in the community with respect to the last data point available. In this scenario we include also a relaxation of two levels (i.e., subtracting 2, if possible) to the Oxford Coronavirus Government Response index on week 6, 2021 [11].

In Figure 4 we show the evolution over time of the leading eigenvalue of the contacts matrix for different basins under different scenarios. This quantity is important as it is one of the contributions to the reproductive number of the model. For comparison, we also report the eigenvalue of the baseline contacts matrix with no restrictions (grey dashed horizontal line). We observe that, while the proposed scenarios correspond to an increase in the eigenvalue and thus in the effective reproductive number, the value corresponding to an unconstrained, pre-pandemic, scenario is much higher.

**Figure 4:**
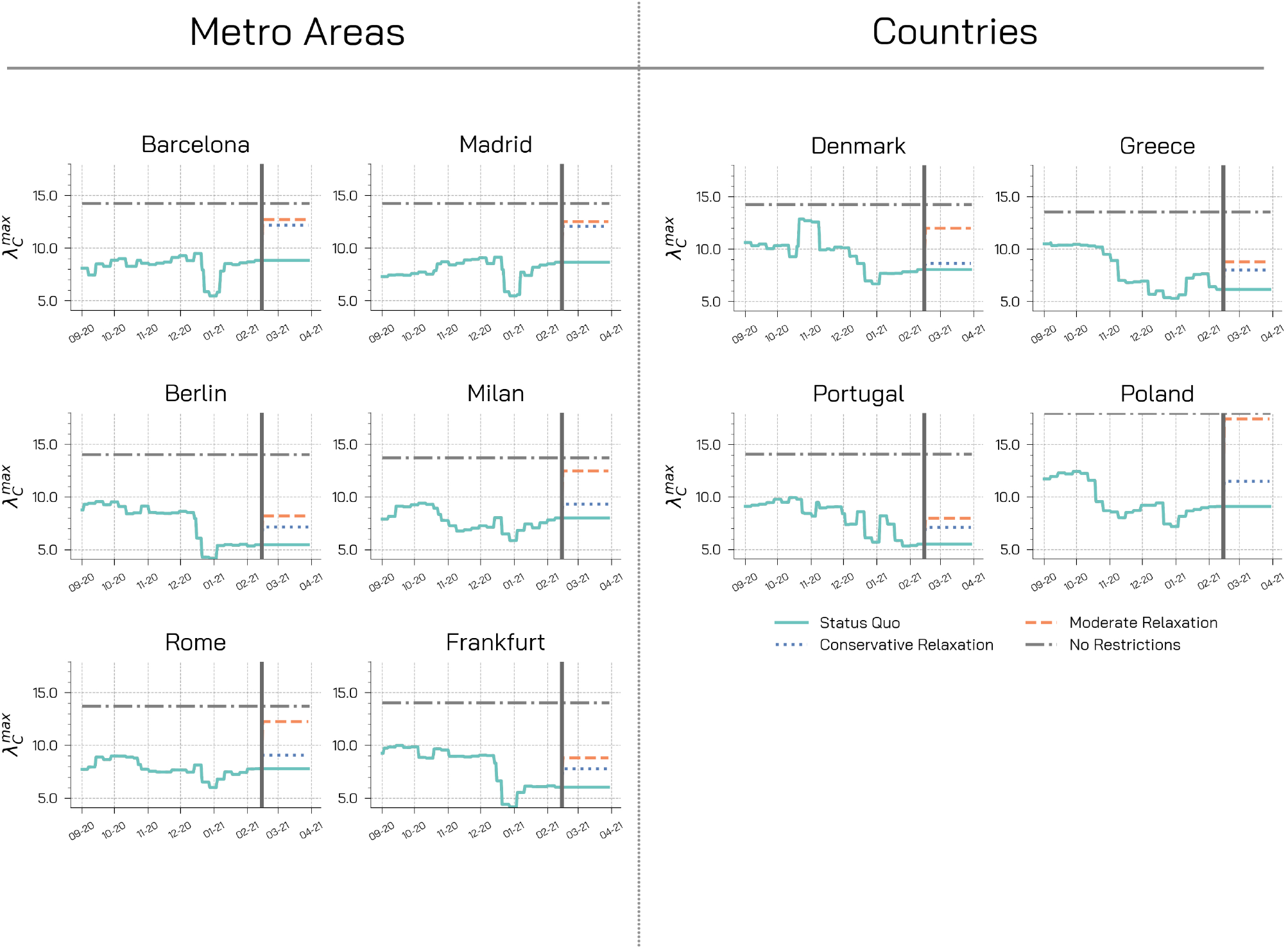
Maximum eigenvalue of the contact matrices. We represent the maximum eigenvalue of the overall contacts matrix over time. The vertical line indicates where the restriction scenarios start. We consider three possible scenarios. In the first scenario (`status quo`), we keep contact matrices unchanged with respect to week 6 of 2021. We then propose two scenarios describing a conservative and a moderate relaxation of social distancing and other NPIs.

### Modeling B.1.1.7 variant introductions

In order to simulate the introductions of B.1.1.7 variant infections in each geographical area we use GLEAM, a global stochastic metapopulation model that simulates the mobility of people across more than 3,300 sub-populations in about 190 countries/territories[7, 8, 33]. Sub-populations are defined by the catchment area of major transportation hubs. The mobility among sub-populations integrates both the long-range the mobility from global air travel (obtained from the International Air Transport Association and Official Airline Guide (OAG) databases) and the short-scale mobility between adjacent sub-populations, which represents the daily commuting patterns of individuals. For international airline travel we use year 2020 data on passengers (origin-destination data) provided by the OAG [34]. The model is calibrated to the initial international importation of cases in the early phases of the pandemic from China and the evolution of deaths in each country. It also considers the set of travel restrictions, mobility reductions, and government interventions. In order to take into account the stochastic nature of introductions of the variant and the onset of local transmission, we consider 307, 000 stochastic runs generated by the model. In particular, we consider only arrivals of individuals in the latent compartment for each age bracket. Indeed, travelers from foreign destinations are now required to exhibit a negative test and other checks are conducted at the airports to avoid symptomatic individuals to travel. The first two specimens of the B.1.1.7 variant were collected on September 20 and 21, 2020 in London and Kent areas, respectively. As UK sequences about 5% of positive cases [35], we modeled the emergence of the B.1.1.7 variant on week 38 of 2020 assuming a cluster of symptomatic/exposed infectious individuals drawn from a Poisson distribution with mean value of 40 symptomatic individuals. We set *ψ* = 0.5, hence assuming the new variant as 50% more transmissible. Below we report a sensitivity analysis for *ψ* = 0.3 and *ψ* = 0.7.

### Model Calibration

We start the simulations on 2020/09/01. The initial distribution of individuals in *S, L*_1_, *I*_1_, and *R* compartments, in each area, is obtained from GLEAM calibrated to international importations of cases from China in the early phases of the pandemic and confirmed deaths profiles in each country. In each administrative regions considered here we assign individuals to each compartment according to the share of cumulative prevalence in the region from real epidemiological data. For example, if a region reported 20% of all cases in the country, we will assign to the *R* compartment of this region 20% of GLEAM country projections. Similarly, we assign individuals in *I*_1_, *E*_1_ compartments according to the share of last week incidence in the region. We use a latent period *ϵ* ^−1^ of 4 days and infectious period of 2.5 days (which implies a generation time *T*_*G*_ of 6.5 days), in line with current estimates [36,37].

The model is calibrated over the period 2020/09/01 − 2021/02/14 using an Approximate Bayesian Computation technique [9]. Free parameters are sampled from a prior distribution and an instance of the model is generated for these parameters. An output quantity of the model *E*^*’*^ is compared to the real quantity *E* using a distance metric *S*(*E, E*^*’*^). If this distance is smaller (greater) than a predefined tolerance *ϵ*, than the sampled parameters set is accepted (discarded). This procedure is repeated iteratively until *N* sets are accepted. The distribution of the accepted sets will approximate the real posterior distribution. Here, we consider weekly reported deaths as output quantity, weighted mean absolute percentage error as distance metric (with tolerance *ϵ* = 0.35), and *N* = 10, 000. We set uniform priors on the the effective reproduction number *R*_*t*_ *∼* [0.9, 2.0] (on 2020/09/01), on the seasonality parameter *α* ∼ [0.5, 1.0], and on the delay in deaths Δ ∼ [12, 25]. Indeed, for COVID-19 the average time between symptoms onset and death is about 2 weeks [29] and we also account for possible additional delays in death reporting. We select also the initial conditions considering the estimates provided by GLEAM on 2020/09/01. We then generate model projections from an ensemble of 5, 000 possible trajectories sampled from the posterior distribution.

In Fig. 5 we represent the obtained posterior distributions for *R*_*t*_ on 2020/09/01, while in Fig. 6 we represent the posterior distributions for Δ, *α*_*min*_, and the initial conditions. In Figure 7 we show the model adequacy after calibration for each region under investigation representing projected and real weekly deaths. Overall, the model is able to capture the temporal patterns observed in the weekly number of reported deaths with nearly all data falling with 95% of model predictions. It is important to remark that the data refers to the number of reported deaths by date of reporting (instead of by date of deaths); as such, it suffers of the typical bias of reporting systems. This may partially explain the high large weekly fluctuations observed in the data for Barcelona.

**Figure 5:**
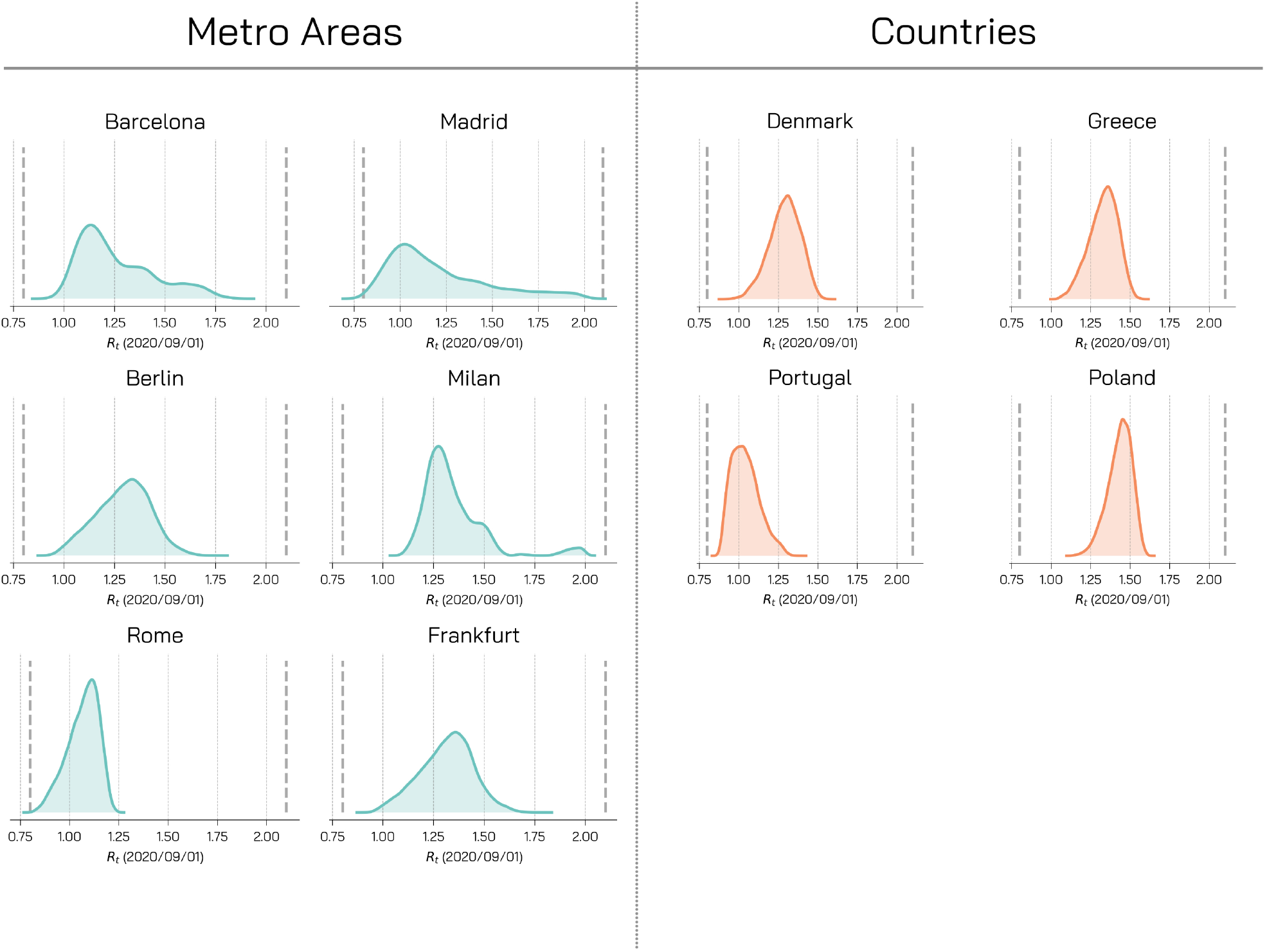
Posterior Distributions - *R*_*t*_. For different regions, we represent the posterior distributions for *R*_*t*_ on 2020/09/01. These posterior distributions are obtained for *ψ* = 0.5 and weighted mean absolute percentage error as a distance metric with a tolerance *ϵ* of 0.35.

**Figure 6:**
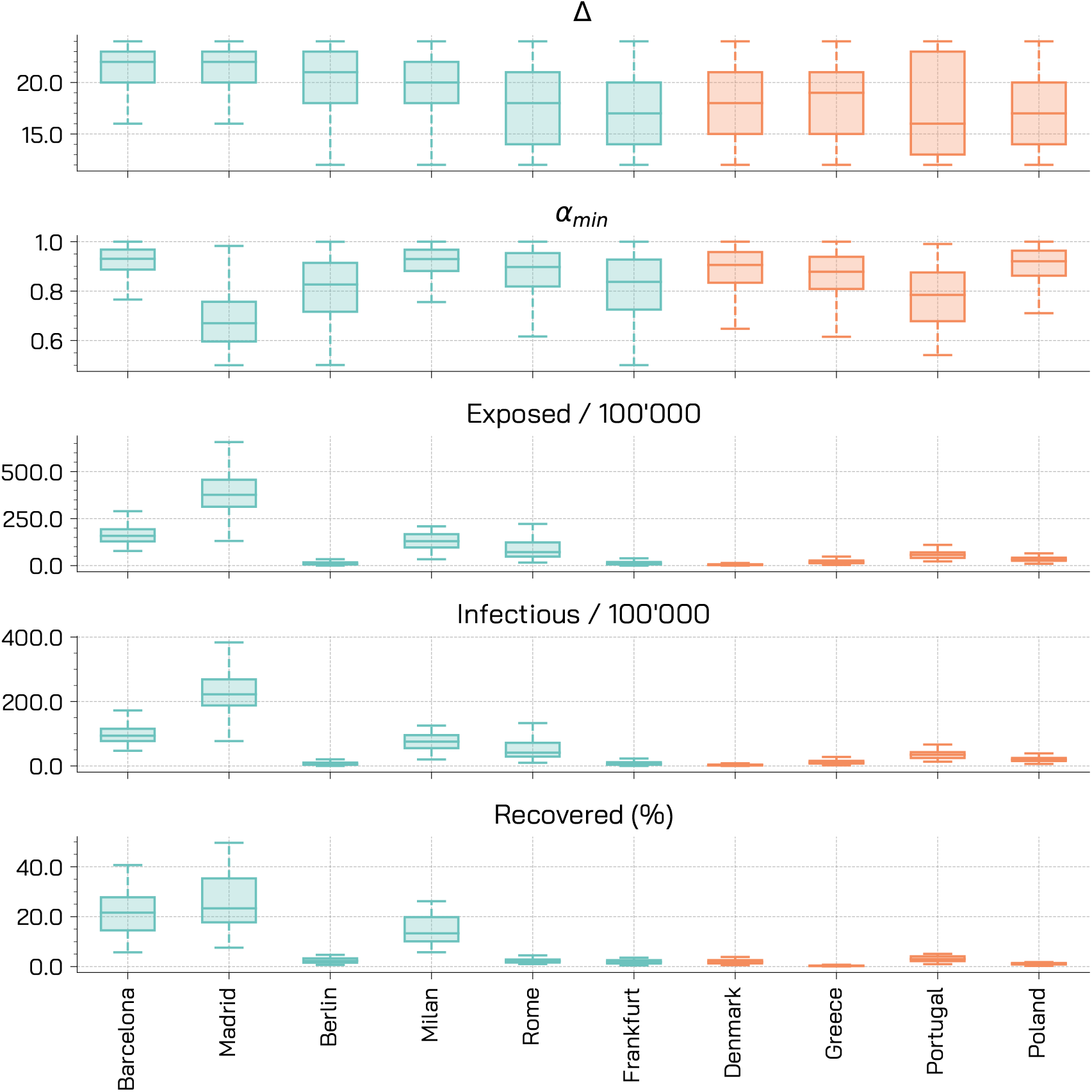
Posterior Distributions - Other parameters. For different regions, we represent the posterior distributions for the delay in deaths Δ, the seasonality parameter *α*_*min*_, and the initial conditions expressed as total number of initial exposed and infectious per 100^’^000 and the percentage of initial recovered. These posterior distributions are obtained for *ψ* = 0.5 and weighted mean absolute percentage error as a distance metric with a tolerance *ϵ* of 0.35.

**Figure 7:**
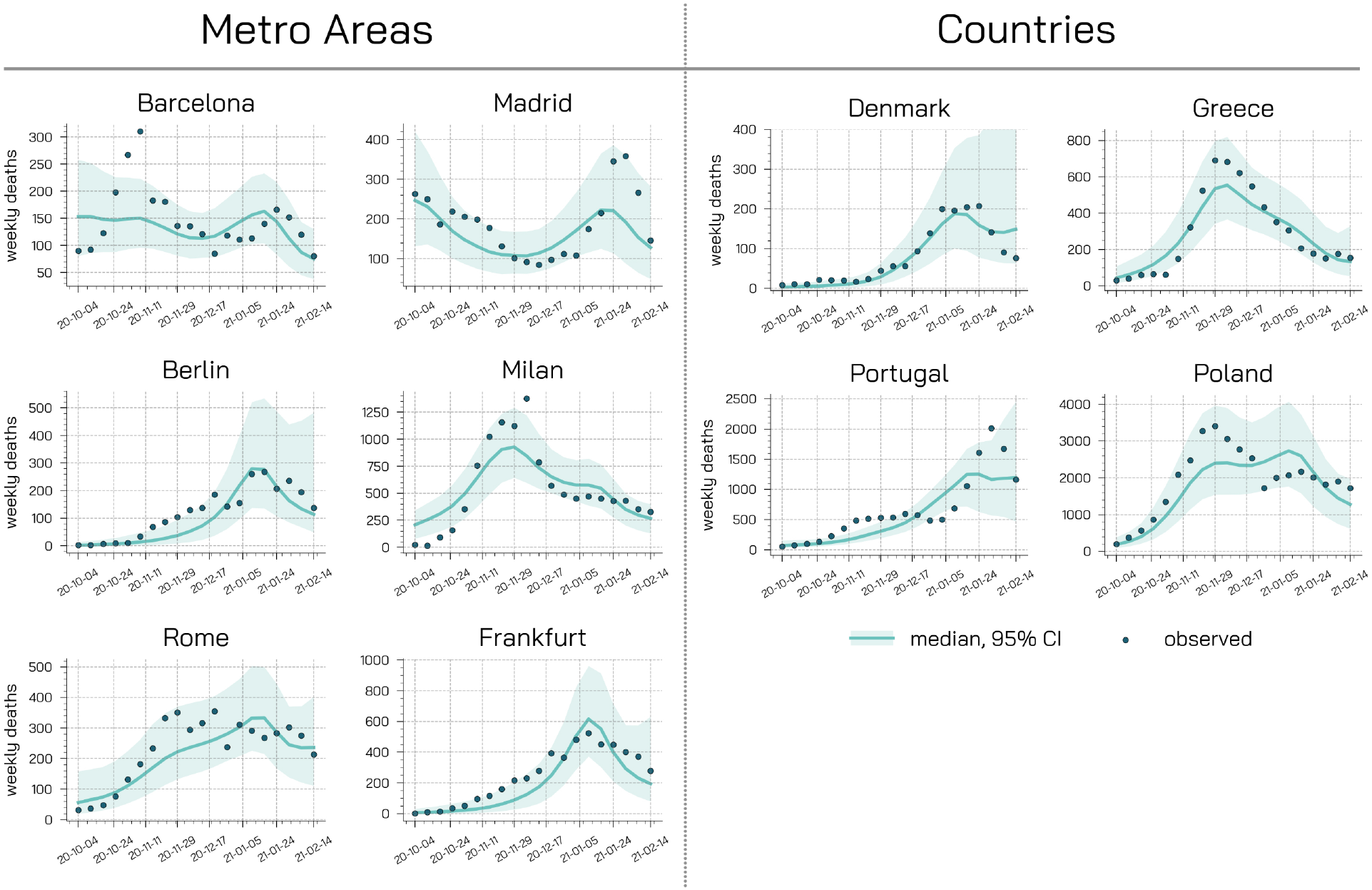
Model adequacy provided by comparing the weekly deaths data with the projections of the calibrated model.

### Effective reproductive number

In Figure 8 we represent the evolution of the effective reproductive number *R*_*t*_ in the regions considered under the different restriction scenarios. We observe that the introduction of the new variant causes an increase in the reproductive number across the board. It is worth noticing that also in the `status quo` scenario (i.e., baseline), *R*_*t*_ is significantly affected pushing the value gradually above the critical threshold *R*_*t*_ = 1. Not surprisingly, higher and faster increase are observed when mitigation measures are relaxed. These observations paint a very concerning and sobering picture. In fact, in case the B.1.1.7 variant would be able to spread, current non-pharmaceutical interventions might not be enough to control it. In other words, our observations hint to possible third waves similarly to what has happened in the UK and it is currently happening in Portugal.

**Figure 8:**
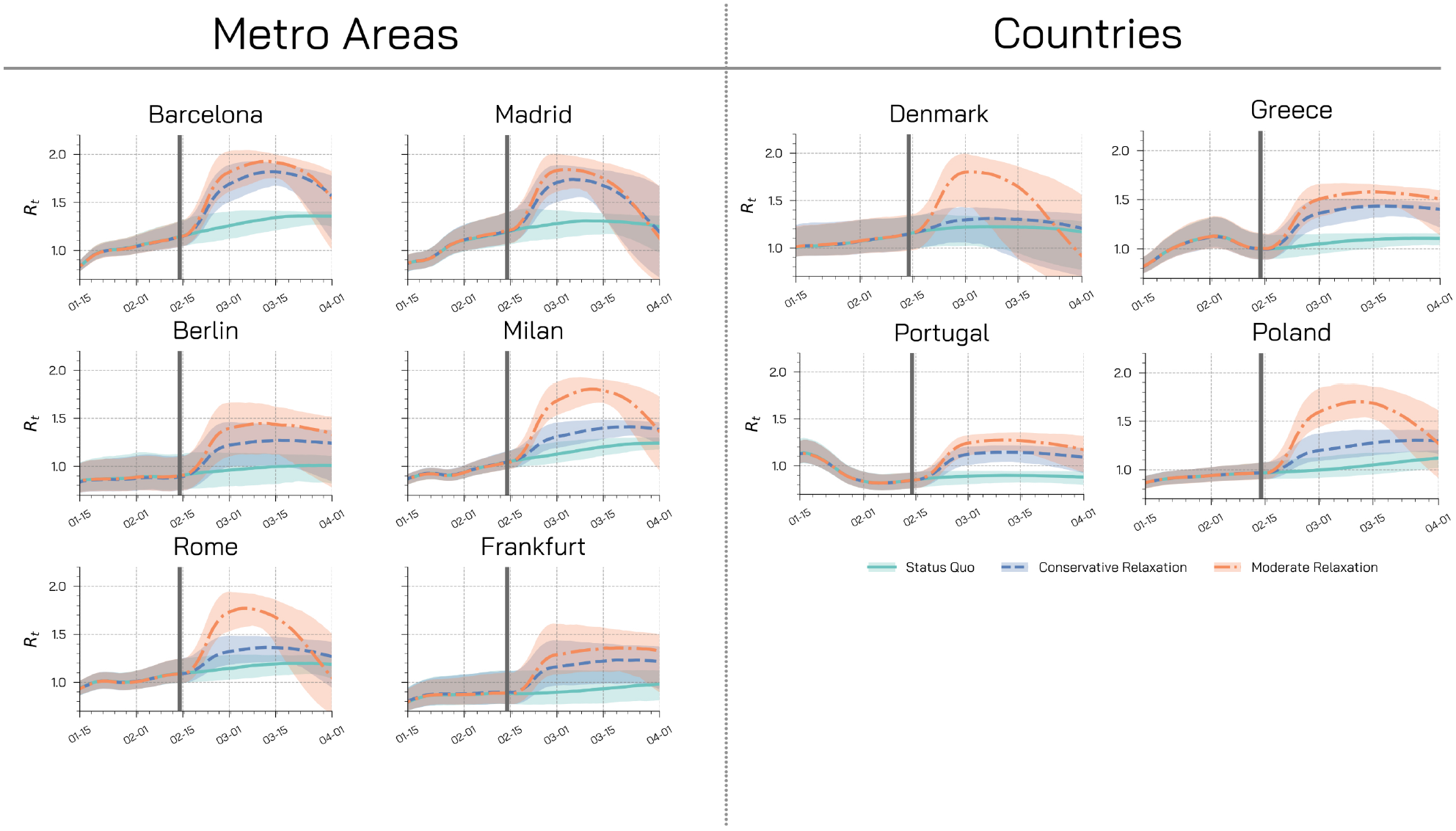
Effective Reproductive Number. Estimate of *R*_*t*_ (median and 90% CI) in different regions for different restrictions scenarios. The B.1.1.7 variant is assumed to have a 50% increase in transmissibility.

### Sensitivity analysis

In the main text we considered an increased transmissibility of VOC 202012/01 of 50%, here we present results also for 30% and 70%. We repeat the calibration step with *ψ* = 0.3 and *ψ* = 0.7. In Fig. 9, 10, 12, 13, we observe that the posterior distributions of the free parameters are not significantly affected by different *ψ*. Similarly, also the projected weekly deaths in Fig. 11, 14 are compatible with those obtained for *ψ* = 0.5. In Fig 15 and 16 we represent the evolution in time of the fraction of new cases attributable to VOC 202012/01 for, respectively, *ψ* = 0.3 (transmissibility increased by 30%) and *ψ* = 0.7 (transmissibility increased by 70%). As expected, for the lower value of *ψ* the growth of VOC 202012/01 is much more contained over time (in most of the cases it does not become dominant by end of March), while for the higher value the growth is much more consistent (and the dominance date is anticipated of about 2 weeks across the board).

**Figure 9:**
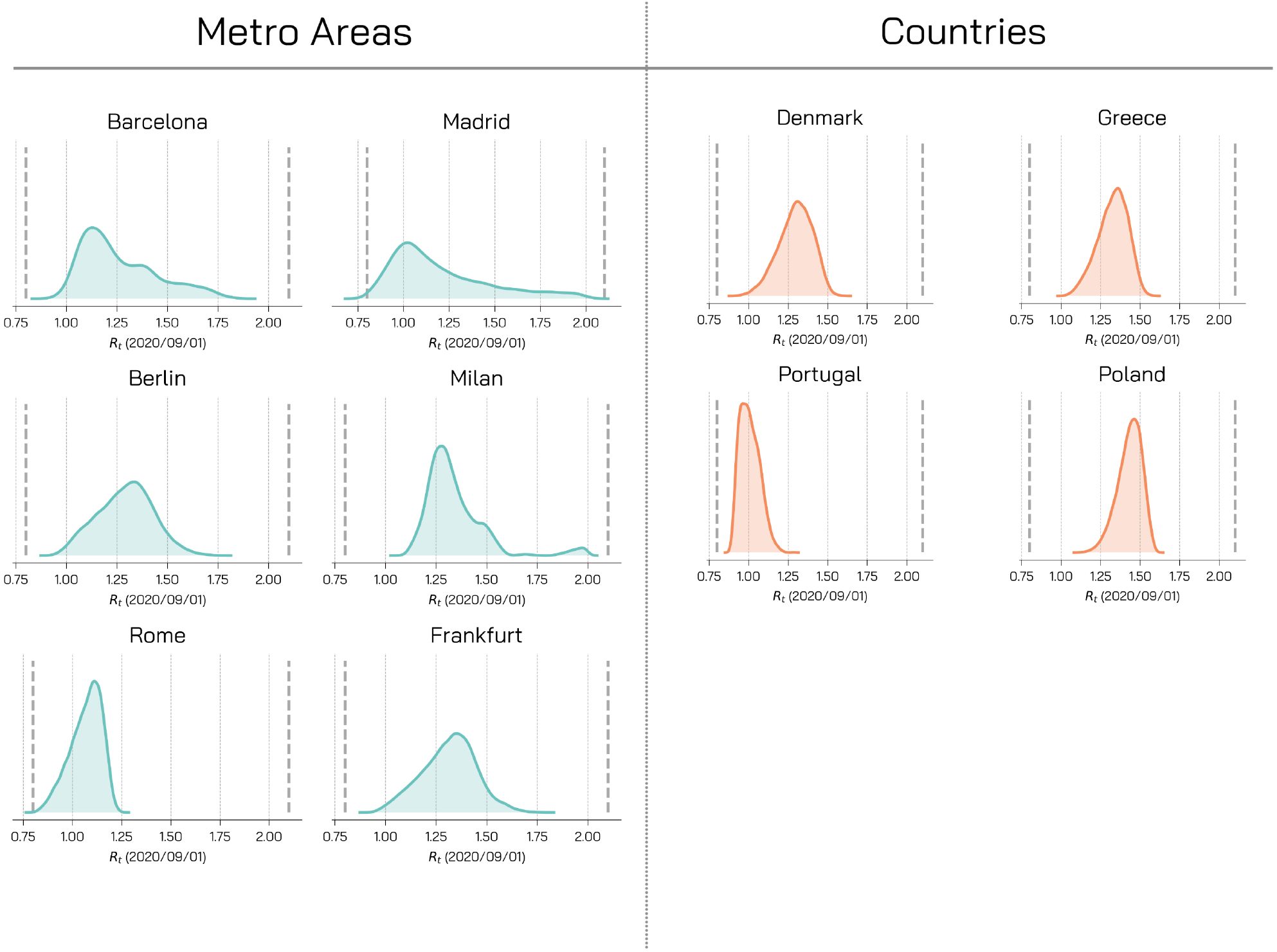
Posterior Distributions - *R*_*t*_. For different regions, we represent the posterior distributions for *R*_*t*_ on 2020/09/01. These posterior distributions are obtained for *ψ* = 0.3 and weighted mean absolute percentage error as a distance metric with a tolerance *ϵ* of 0.35.

**Figure 10.**
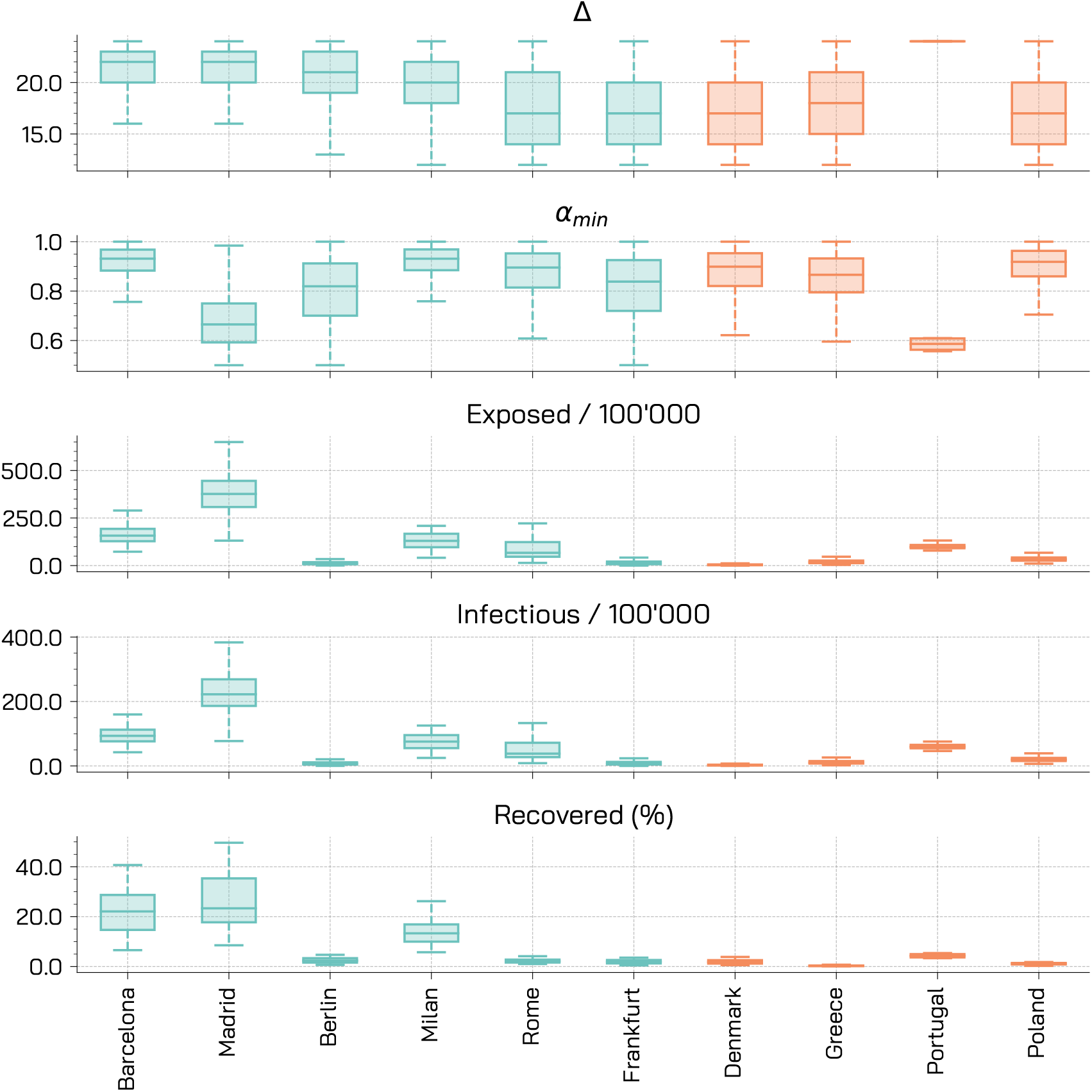
Posterior Distributions - Other parameters. For different regions, we represent the posterior distributions for the delay in de deaths Δ, the seasonality parameter *α*_*min*_, and the initial conditions expressed as total number of initial exposed and infectious per 1000’000 and the percentage of initial recovered. These posterior distributions are obtained for *ψ* = 0.3 and weighted mean absolute percentage error as a distance metric with a tolerance *ϵ* of 0:35.

**Figure 11:**
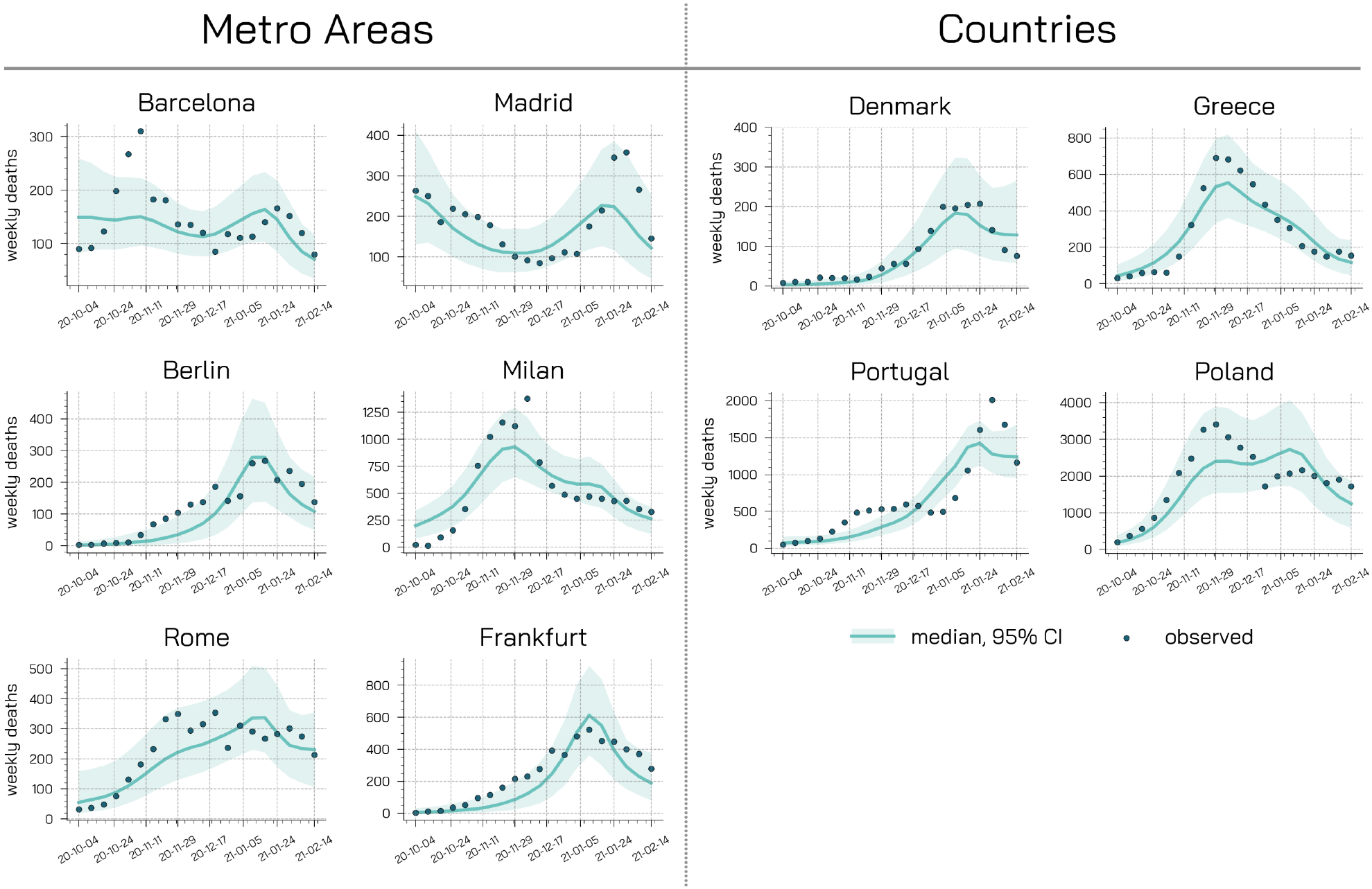
Model adequacy provided by comparing the weekly deaths data with the projections of the calibrated model (*ψ* = 0.3).

**Figure 12:**
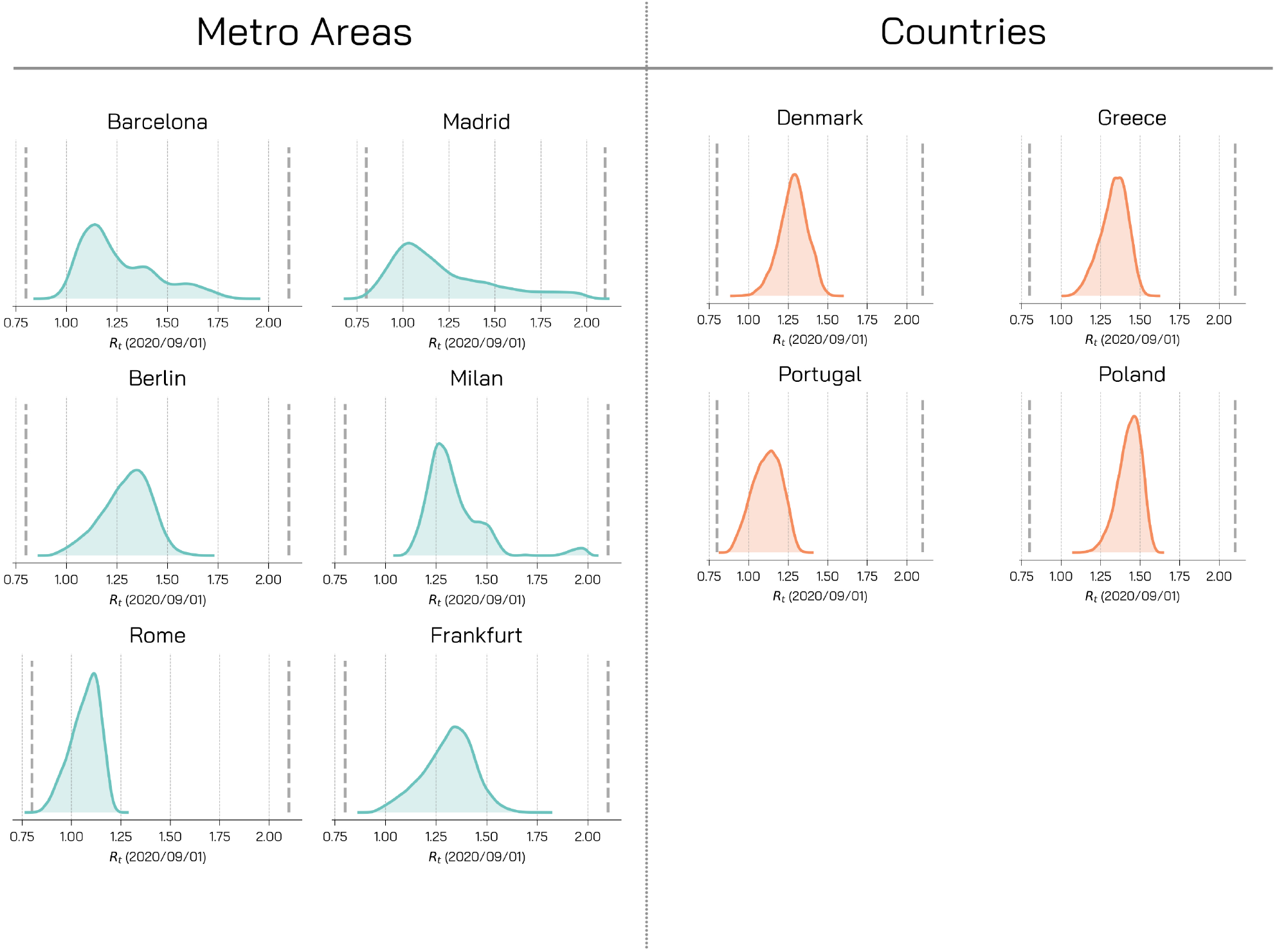
Posterior Distributions - *R*_*t*_. For different regions, we represent the posterior distributions for *R*_*t*_ on 2020/09/01. These posterior distributions are obtained for *ψ* = 0.7 and weighted mean absolute percentage error as a distance metric with a tolerance *E* of 0.35.

**Figure 13:**
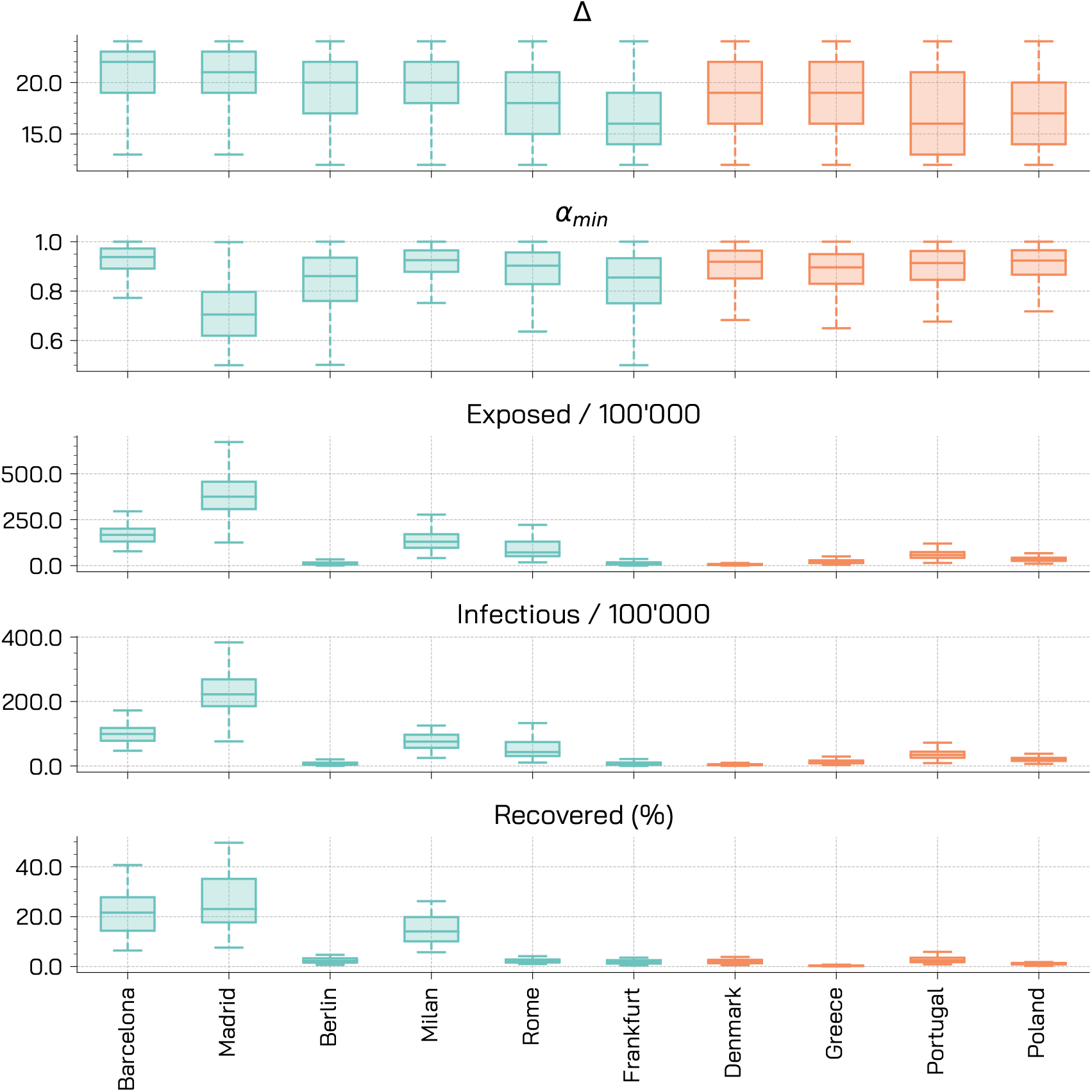
Posterior Distributions - Other parameters. For different regions, we represent the posterior distributions for the delay in de deaths Δ, the seasonality parameter *α*_*min*_, and the initial conditions expressed as total number of initial exposed and infectious per 100^’^ 000 and the percentage of initial recovered. These posterior distributions are obtained for *ψ* = 0.7 and weighted mean absolute percentage error as a distance metric with a tolerance *ϵ* of 0.35.

**Figure 14:**
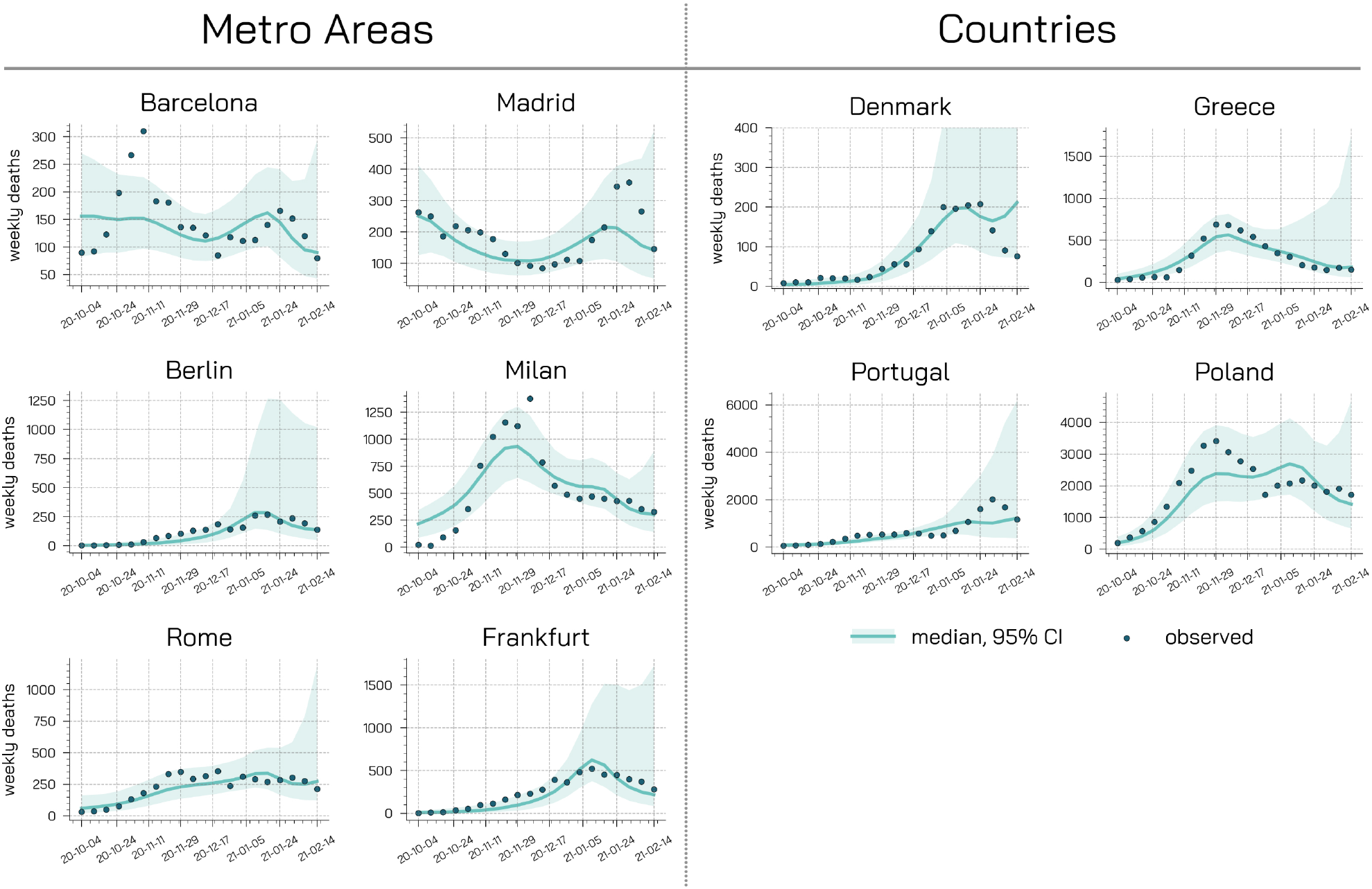
Model adequacy provided by comparing the weekly deaths data with the projections of the calibrated model (*ψ* = 0.7).

**Figure 15:**
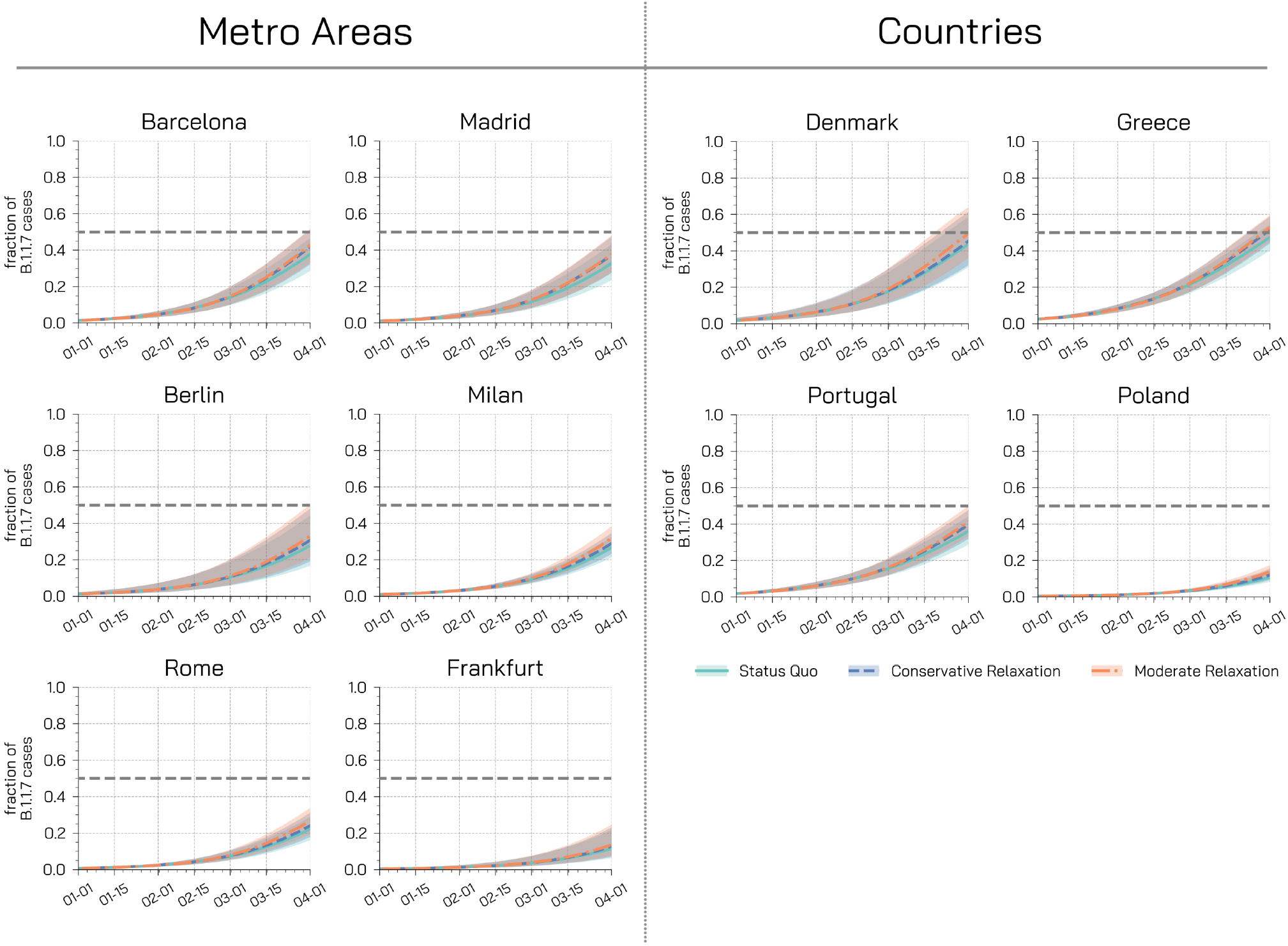
Fraction of new cases attributable to the B.1.1.7 variant. Each plot shows the fraction of new weekly cases attributable to the variant in different regions for different restrictions scenarios under the assumption of a 30% increase in transmissibility. Dashed horizontal lines represent the dominance threshold of 50% of new cases caused by the B.1.1.7 variant. The shaded areas represent the 50% CI.

**Figure 16:**
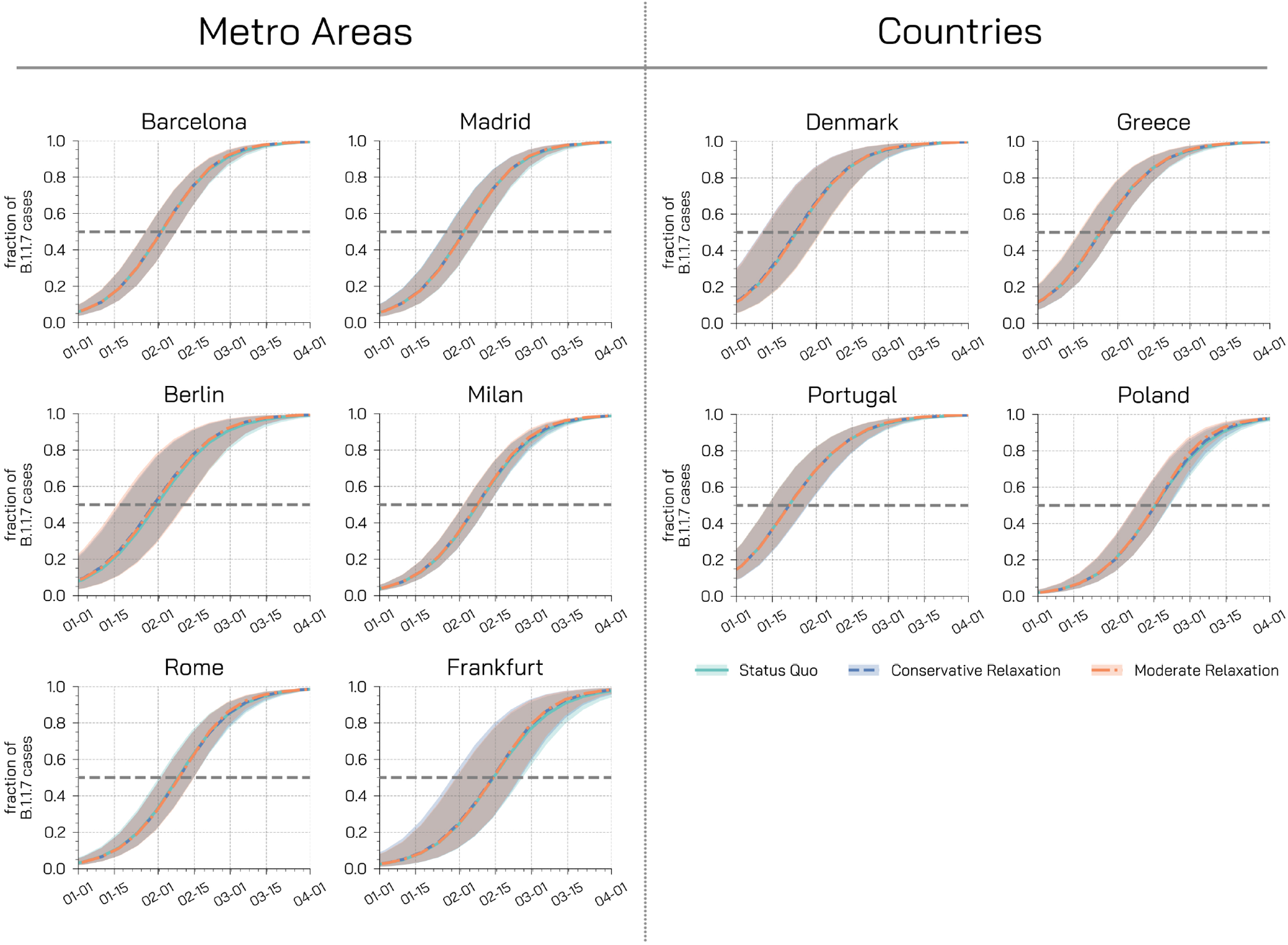
Fraction of new cases attributable to the B.1.1.7 variant. Each plot shows the fraction of new weekly cases attributable to the variant in different regions for different restrictions scenarios under the assumption of a 70% increase in transmissibility. Dashed horizontal lines represent the dominance threshold of 50% of new cases caused by the B.1.1.7 variant. The shaded areas represent the 50% CI.

## Notes

### Competing Interest Statement

A.V. reports grants and personal fees from Metabiota inc., outside the submitted work; M.C. reports grants from Metabiota inc., outside the submitted work. M.A. reports research funding from Seqirus; the funding is not related to COVID-19. No other relationships or activities that could appear to have influenced the submitted work.

### Funding Statement

M.C., M.A., and A.V. acknowledge support from COVID Supplement CDC-HHS-6U01IP001137-01. M.C. and A.V. acknowledge support from Google Cloud and Google Cloud Research Credits program to fund this project. The findings and conclusions in this study are those of the authors and do not necessarily represent the official position of the funding agencies, the National Institutes of Health, or the U.S. Department of Health and Human Services.

